# Defining high-value information for COVID-19 decision-making

**DOI:** 10.1101/2020.04.06.20052506

**Authors:** COVID-19 Statistics, Policy modeling and Epidemiology Collective (C-SPEC), Alyssa Bilinski, Ruthie Birger, Samantha Burn, Melanie Chitwood, Emma Clarke-Deelder, Tyler Copple, Jeffrey Eaton, Hanna Ehrlich, Margret Erlendsdottir, Soheil Eshghi, Monica Farid, Meagan Fitzpatrick, John Giardina, Gregg Gonsalves, Yuli Lily Hsieh, Suzan Iloglu, Yu-Han Kao, Evan MacKay, Nick Menzies, Bianca Mulaney, David Paltiel, Stephanie Perniciaro, Maile Phillips, Katherine Rich, Joshua A Salomon, Raphael Sherak, Kayoko Shioda, Nicole Swartwood, Christian Testa, Thomas Thornhill, Elizabeth White, Anne Williamson, Anna York, Jinyi Zhu, Lin Zhu

## Abstract

Initial projections from the first generation of COVID-19 models focused public attention on worst-case scenarios in the absence of decisive policy action. These underscored the imperative for strong and immediate measures to slow the spread of infection. In the coming weeks, however, as policymakers continue enlisting models to inform decisions on COVID-19, answers to the most difficult and pressing policy questions will be much more sensitive to underlying uncertainties. In this study, we demonstrate a model-based approach to assessing the potential value of reducing critical uncertainties most salient to COVID-19 decision-making and discuss priorities for acquiring new data to reduce these uncertainties. We demonstrate how information about the impact of non-pharmaceutical interventions could narrow prediction intervals around hospitalizations over the next few weeks, while information about the prevalence of undetected cases could narrow prediction intervals around the timing and height of the peak of the epidemic.

## Introduction

The novel coronavirus SARS-CoV-2 has spread rapidly around the world since its December 2019 identification in Wuhan, China.^1,2^ Cases of COVID-19, the illness caused by the virus, have been diagnosed in all 50 United States and are rising sharply.^3^ Many school districts, counties, and states have adopted strong measures to promote social distancing, restrict movement, and reduce transmission.^4,5^ Decision makers have been compelled to set policies based on a rapidly evolving but limited evidence base, and this challenge has been met with a proliferation of epidemiological modeling efforts.

The first generation of models generated forecasts that focused public attention on worst-case scenarios in the absence of decisive policy action. Despite substantial uncertainties surrounding the biology of the disease and intervention costs and effects, most models arrived at the same qualitative finding: left unchecked, the spread of COVID-19 would overwhelm hospital capacity to care for acute and critical cases, resulting in intolerable potential morbidity and mortality. These projections underscored the imperative for bold and immediate measures to slow the spread of infection.^6–8^

Policymakers will continue enlisting models to forecast the trajectory of COVID-19 in the coming weeks.^6,9,10^ As many communities have implemented shelter-in-place orders and other non-pharmaceutical interventions (NPIs), vital questions now include how to plan for short-term and long-term health system needs and when and how policymakers might begin to modify or relax the imposition of NPIs. Answers to these questions will be much more sensitive to underlying uncertainties. They will require a different kind of modeling, focused less on conventional forecasting and more on program evaluation, resource allocation, careful characterization of uncertainty, and optimal inference from limited information.

In this study we demonstrate a model-based approach to determining priorities for acquiring new information based on the potential value of this information in reducing critical uncertainties relevant to COVID-19 decision-making over short- and medium-term. We fit an age-structured compartmental model to reported cases in the San Francisco Bay Area, forecast a range of possible epidemic trajectories consistent with available information, and then demonstrate how additional information about the impact of NPIs or the fraction of all infections that are confirmed positive would impact case predictions.

## Methods

We developed an age-structured dynamic compartmental model of COVID-19 transmission, similar to previously published models.^10^ Model compartments stratify the population into susceptible, exposed, symptomatic infected, asymptomatic infected, and recovered individuals. We derived initial ranges for model parameters including length of incubation and infectious periods, and the basic reproductive number (R0), by drawing from prior COVID-19 modeling studies (**Table 1)**, and from prior studies on age-specific contact patterns (sources in **Supplemental Information**). Initial parameter ranges were specified to envelop at least the high and low point estimates used in other modeling studies, and to encompass uncertainty ranges from those studies when provided (sources in **Supplemental Information**).

**Table 1.** Parameter ranges

| Parameter | Range |
| --- | --- |
| Basic reproductive number | (2, 3) |
| Relative infectiousness given asymptomatic | (.25, .66) |
| Proportion of cases that are asymptomatic |  |
| Ages <20y | (.5, .9) |
| Ages 20-64y | (.2, .4) |
| Ages 65+ | (.2, .4) |
| Average duration of incubation period | (4-6) |
| Average duration of infectious period | (3-7) |
| Percentage of cases ascertained as of March 15 | (4-30) |

Using a Bayesian calibration approach,^11^ we fit model parameters to approximate the pooled population and rise in confirmed COVID-19 cases through March 15 across six counties in the San Francisco Bay Area. These counties collectively comprise an early epicenter of spread in the US and were also early adopters of NPIs. We projected two scenarios from March 16 through June 30: a ‘no intervention’ counterfactual scenario and an ‘NPI scenario’ in which we assumed that all contact rates would be reduced by 10% to 60% after March 15.

We assumed that 33% of ascertained cases required hospitalization,^12^ and back-calculated the proportion of all cases requiring hospitalizations for a given set of parameter values (**Supplemental Information**). We assumed that average duration of hospitalization for COVID-19 was 10 days.^6^ We estimated a hospital capacity constraint using AHA estimates and assumed that 50% of beds could be available for COVID-19 patients.^12,13^

To assess the potential decision value of new information, we focused on the projected time until the hospital supply constraint was exceeded by COVID-19 demand as a proxy for major decision-relevant outcomes. First, we calculated the uncertainty around this projected time when the model was only calibrated to the confirmed case counts through March 15. Second, we evaluated the reduction in this uncertainty that would result from obtaining more precise information about 1) the effectiveness of NPIs or 2) the fraction of all cases ascertained. For illustration, we considered narrowing uncertainty around NPI effectiveness to either a 10-30% average contact reduction or a 40-60% average contact reduction, and narrowing uncertainty around the ascertainment fraction either to 2-10% or 20-30%.

## Results

Our base case results show that an extraordinarily wide range of scenarios would be consistent with the cumulative confirmed cases through March 15. In the no intervention scenario (**Figure 1**), without any interruptions to the natural spread of infection, and allowing for deliberately broad ranges around parameter values, projections through May 31 span from 165,000 to more than 5 million total cases in the Bay Area, comprising an illustrative worst-case hypothetical. Incorporating the potential impact of NPIs since March 15 leads to a dramatic projected reduction in case numbers and hospitalizations against the hypothetical no intervention scenario, and to a substantially lower probability of exceeding hospital capacity over the projected time horizon.

**Figure 1.**
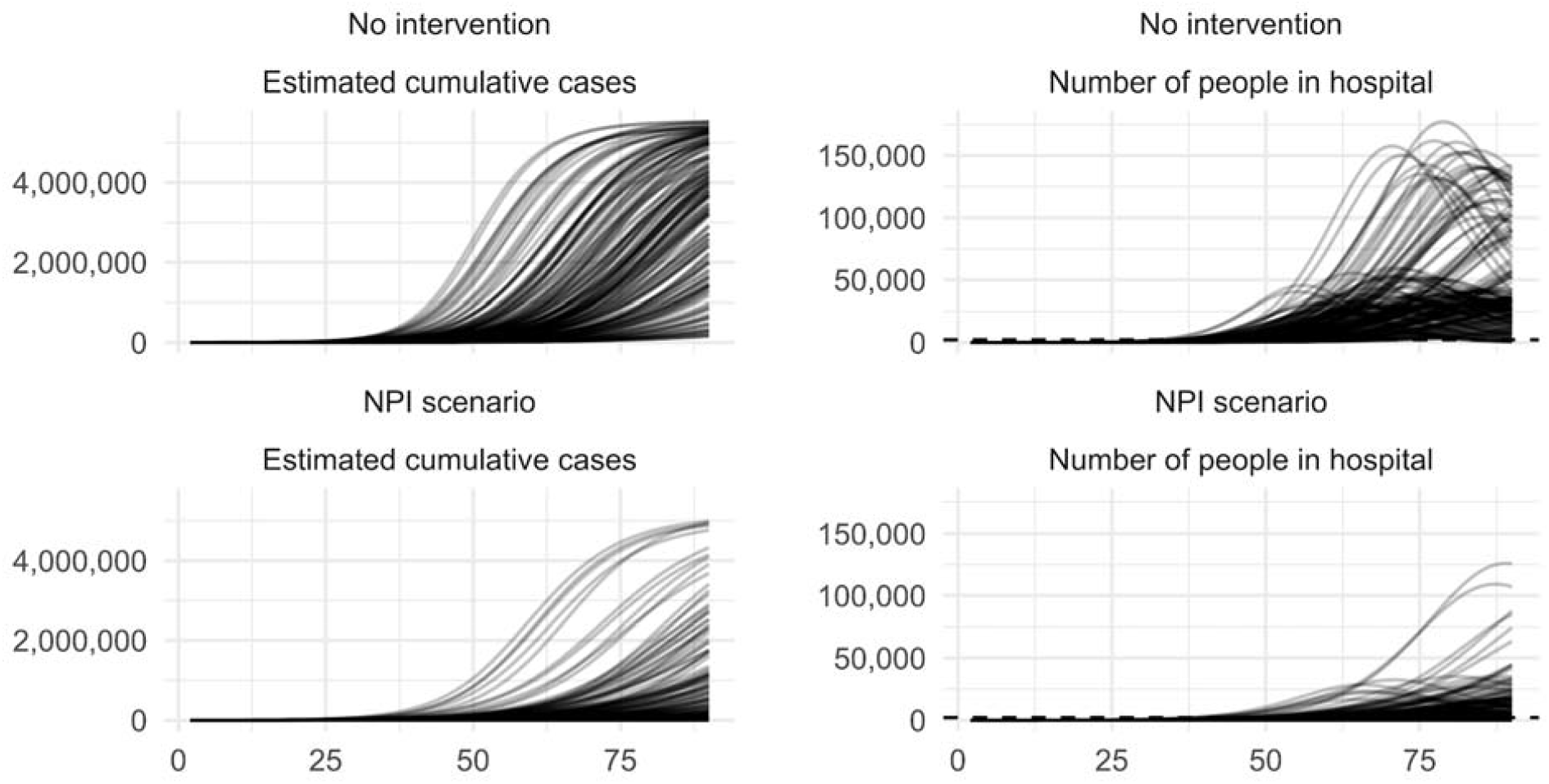
Projected cumulative cases and hospitalizations in no intervention scenario (top row) and NPI scenario (bottom row), assuming no new information is available. Each curve is the trajectory generated by one parameter set in the posterior distribution, fitted to cumulative cases at one time point. The dotted line in the right column indicates estimated hospital capacity.

Without additional information, however, there remains substantial uncertainty around both the no intervention and NPI scenarios. Additional information sources could narrow these ranges considerably. In particular, if new information signalled that social distancing efforts reduce average contacts by 40-60%, the majority of scenarios examined (84%) would not exceed hospital capacity by May 31. If additional information pointed to a relatively low burden of undetected infection (i.e. higher case ascertainment at baseline) (**Figure 2**), this information would signal a longer time to reach the peak of the epidemic and higher peak compared to an alternative in which new information pointed to lower baseline case ascertainment. Combining the two types of new hypothetical information, **Figure 3** shows that knowledge about the impact of social distancing measures would be much more informative for determining when hospital capacity could be reached, relative to the value of additional information about the fraction of cases ascertained.

**Figure 2.**
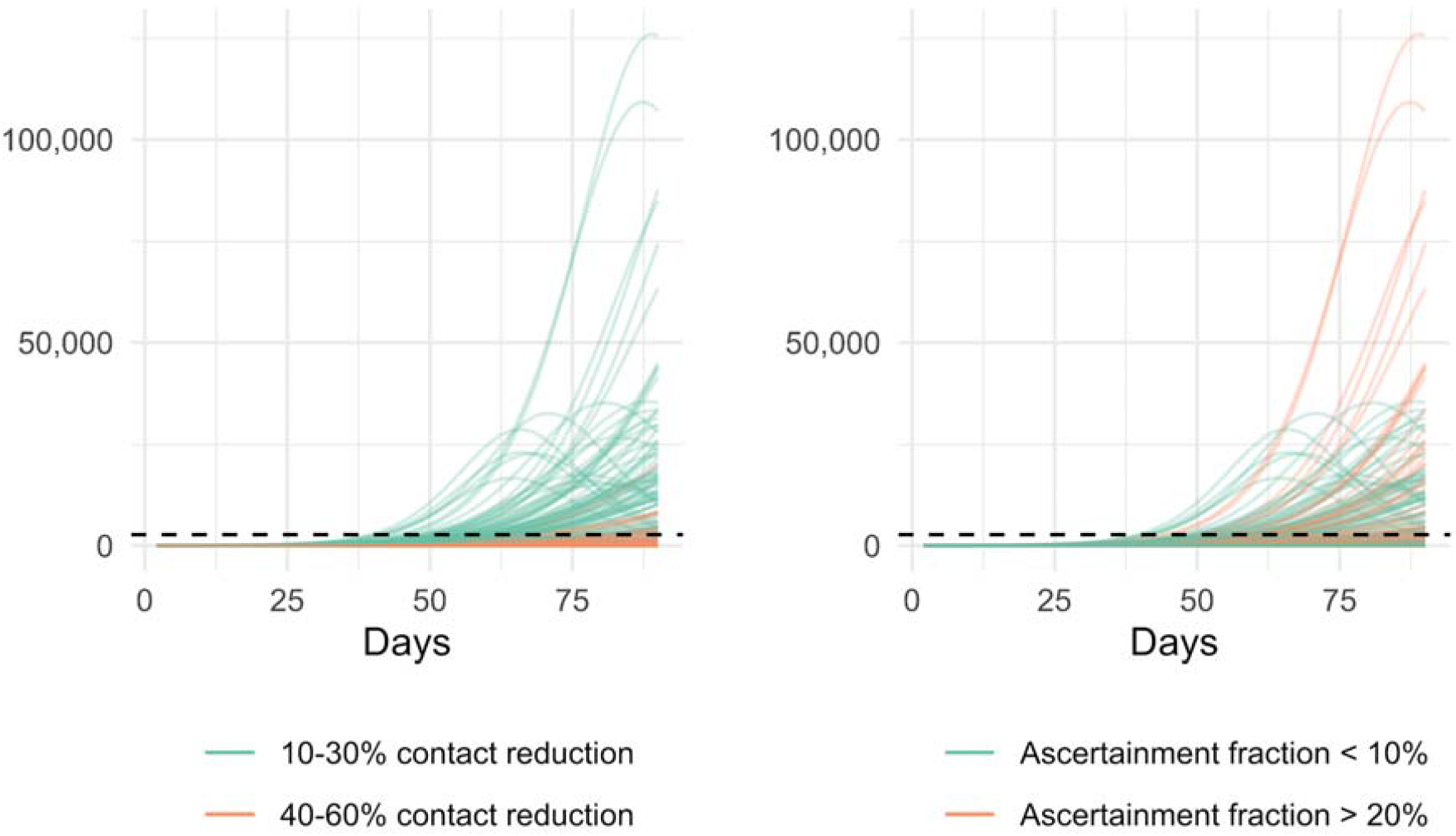
Projected hospitalizations in the NPI scenario assuming hypothetical new information on NPI effects (left panel) or case ascertainment (right panel).

**Figure 3.**
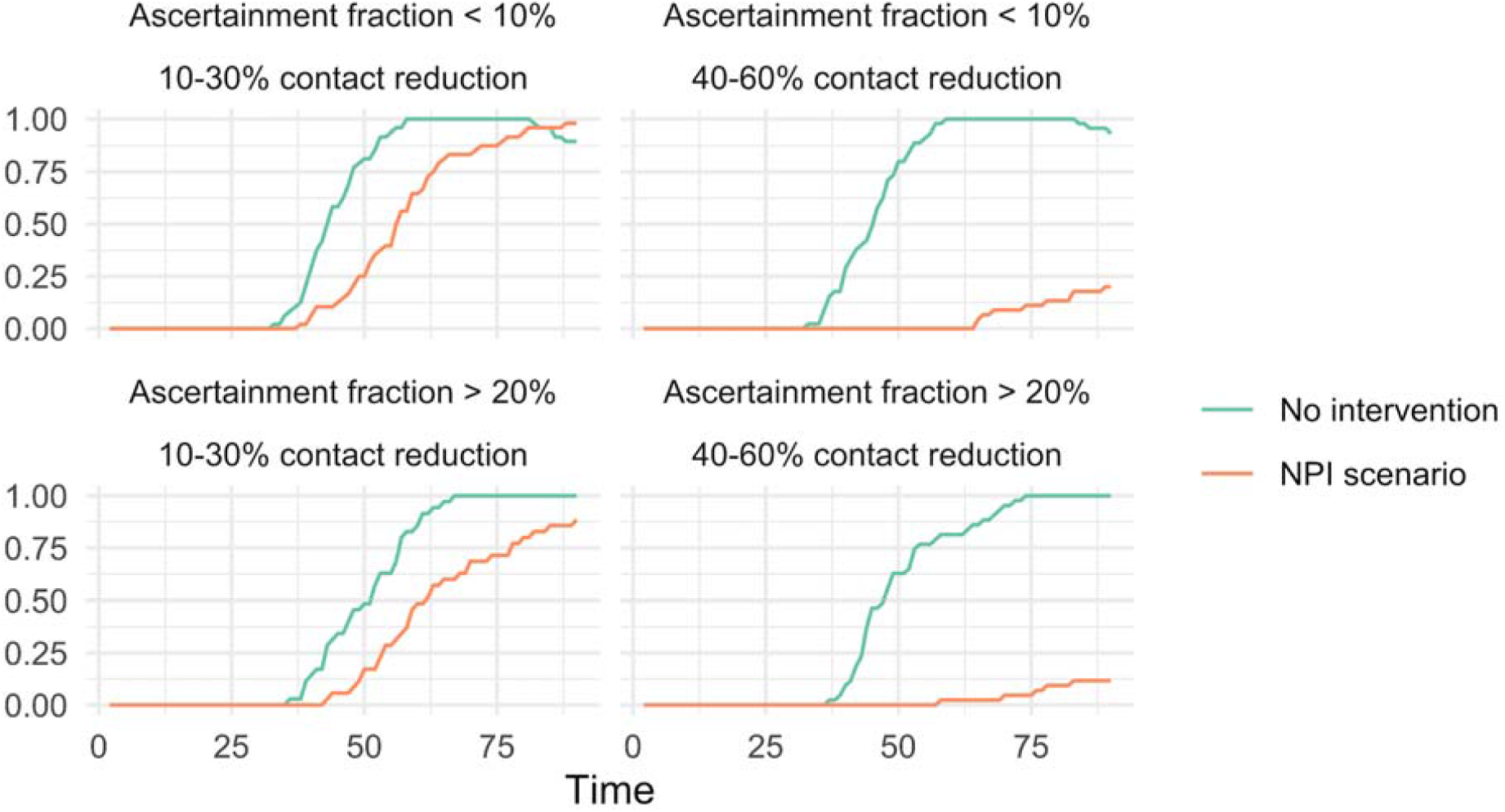
Probability of exceeding hospital bed capacity assuming hypothetical new information on both NPI effects and case ascertainment..

## Discussion

In this paper, we use a simple epidemiological model and a stylized example of how priorities for collecting new information might be evaluated with respect to their value for decision-making in the COVID-19 response. In an environment with considerable uncertainty, it is important to understand the implications that obtaining additional information could have for policy and planning. In the short-term, our results suggest that given baseline knowledge of the number of tests and number of hospitalizations, short-term planning on the scale of weeks would benefit most from understanding the impact of NPIs. Over a longer term, information about the fraction of infections that are not captured in confirmed case reports will be critical to forecasting the extended trajectory of the epidemic, including the timing and level of its peak.

There are numerous limitations to our analysis, including uncertainty around parameter prior distributions and those excluded from the uncertainty analysis for simplicity. Nonetheless, our analysis highlights the benefits of a decision analytic approach for prioritizing the collection of high-value information for policy-making concerning the COVID-19 pandemic.

## Data Availability

Model code is available on GitHub.

## Supplemental Documentation for

### 1 Model

#### 1.1 Model Structure

The model uses the following structure, with susceptible individuals (*S*) progressing to an exposed state (*E*), and then moving to either a symptomatic infected state (*I*) or an asymptomatic state (*A*). Then, individuals in the symptomatic infected state can either recover or die, while individuals in the asymptomatic state can only recover. (Note that *λ* = *βI/N*, discussed below.)

**Figure 1:**
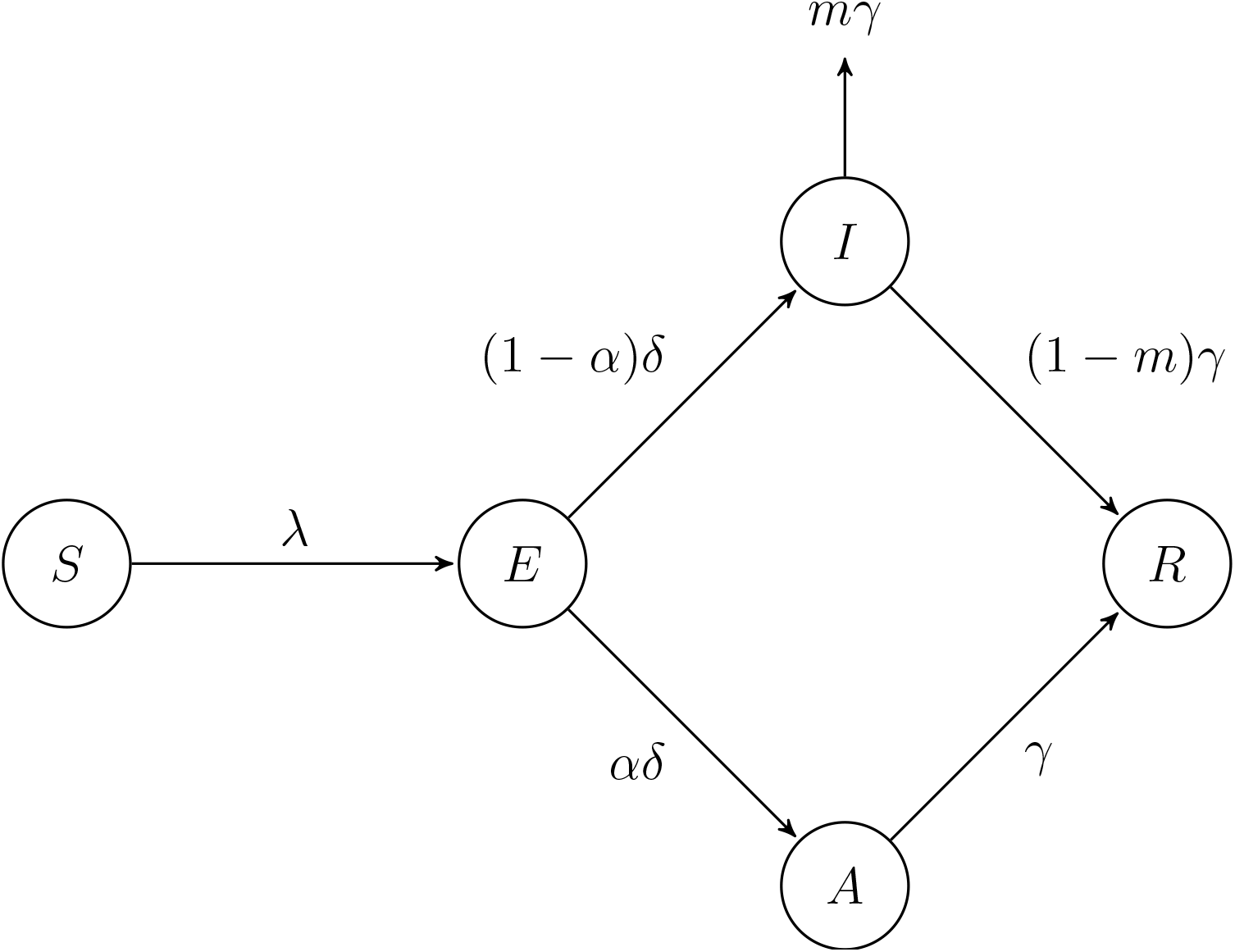
Model Structure

#### 1.2 Baseline Specification

The population is stratified by age into three groups: *i* = {*<* 20, 20 *−* 64, 65+}. Then, let *β*_*ij*_ = *v*_*ij*_*p* be the effective contact rate between susceptible and symptomatic individuals, and let 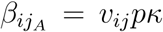 be the effective contact rate between susceptible and asymptomatic individuals. Here, *v*_*ij*_ is the number of daily contacts with individuals in group *j* per person in group *i, p* is the probability of infection per contact between a susceptible and infected individual, and *κ* is the relative reduction in the effective contact rate when the contact is with an asymptomatic individual, compared to a symptomatic individual. Then, the model is defined by the following system of differential equations:

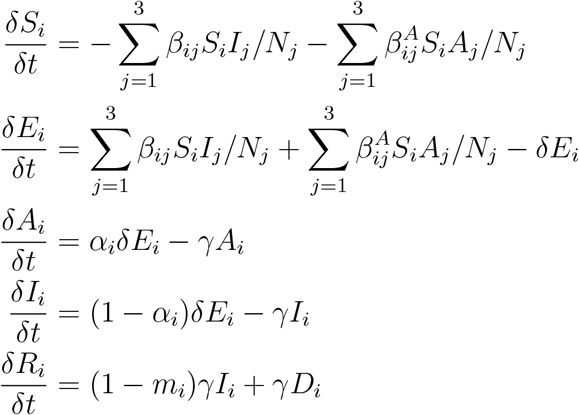

In addition to *β*_*ij*_ defined above, *δ* is the rate of progression out of the exposed state, *α*_*i*_ is the proportion of infections that are asymptomatic for age group *i, γ* is the rate of progression from infected to recovered or death, and *m*_*i*_ is the proportion of individuals in age group *i* who die once they exit the infected state.

#### 1.3 Non-pharmaceutical interventions (NPIs)

In order to model a non-pharmaceutical intervention (NPI) that increases social distancing, the number of contacts across all strata are reduced by a constant fraction – under social distancing, the effective contact rates become 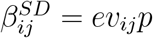 and 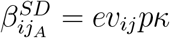, where *e* ∈ [0, 1] (with the remainder of the model identical to the baseline specification). For instance, if *e* = 0.4, the number of contacts will be reduced by 60% across the entire population.

For the analysis in this study, we assumed that the value for *e* (i.e., the effectiveness of the social distancing intervention) is not known with certainty, so the value is drawn from a uniform distribution for each run of the model. The parameters for this uniform distribution were chosen based on the amount of (hypothetical) information available about the effectiveness of social distancing. For the initial scenario, where there is no additional information, *e* is drawn from a range of (0.1, 0.6). For the scenario where additional information is available, *e* is drawn from a range of (0.1, 0.3) or (0.4, 0.6), depending on whether the new information indicated social distancing is relatively effective or ineffective.

### 2 Parameters

#### 2.1 Non-Calibrated Parameters

**Table 1:** Non-Calibrated Parameters

| Parameter | Description | Value |
| --- | --- | --- |
| $N_i(0)$ | Age Distribution at $t = 0$ | $(0.25, 0.59, 0.16)$ |
| $v_{ij}$ | Average number of contacts with individuals in group $j$ per person in group $i$ | (See Below) |
| $m_i$ | Mortality risk for group $i$ | $(0, 0.005, 0.1)$ |

##### 2.1.1 Age distribution

We assume an age distribution representative of six counties in California Bay Area (Alameda, Contra Costa, Marin, San Francisco, San Mateo, and Santa Clara), with 22% of the population under 20, 63% of the population between 20 and 64, and 15% of the population 65 and older (United States Census Bureau, 2019). This is similar to the age distribution in the United States, which has 25% of the population under 20, 59% between 20 and 64, and 16% at least 65 (United States Census Bureau, 2019).

##### 2.1.2 Contact rates between strata

Very limited primary data exists on age-stratified contacts in the US. Therefore, we rely on methods that project contact rates derived from surveys in other contexts and US demographics. We use the age-structured contact matrix for the US estimated by Prem et al. (2017), which was derived by projecting the POLYMOD Mossong et al. (2008, 2017) survey data to US demographics Prem et al. (2017). Since we are interested in 3 broad age-classes, we bin the projected contacts according to US Census Bureau age-distribution estimates for 2018 (United States Census Bureau (2019)). This provided the following baseline contact matrix, where *v*_*ij*_ is the average number of daily contacts with individuals in group *j* per person in group *i* (i.e., *v*_12_ represents the number of daily contacts where an infectious individual in group 2 could infect a susceptible individual in group 1):

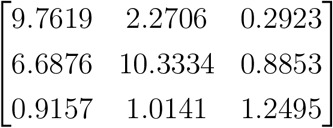

##### 2.1.3 Age-specific mortality rates

We assume that the mortality risks among symptomatic individuals are 0%, 0.4%, and 5.4% for the 0-19, 20-64, and 65+ age groups, respectively. These values were derived from the age-stratified infection fatality ratios (IFR) calculated by Ferguson et al. (2020) (who in turn derived their estimates from work by Verity et al. (2020), while also adjusting for a varying attack rate across age groups). Ferguson et al. (2020) provides an IFR for 10-year age bins (i.e., 0-9 years, 10-19 years, etc.). In order estimate the mortality risk for the three age groups used in this analysis, we calculated a weighted average of the IFR across the bins contained in each group, using estimates for the 2018 US age distribution from United States Census Bureau (2019). (Since the 65 year cutoff used in this analysis occurs in the middle of the 60-69 age bin reported in Ferguson et al. (2020), we linearly interpolated the IFR for the 60-64 and 65-69 age bins.)

It is important to note, however, that the mortality risk values we assume here are somewhat different from other estimates of the mortality risk. Riou et al. (2020), which adjusts crude fatality rates from Hubei province, China, to account for delayed mortality and unidentified cases, estimate that the case fatality risk is less than 0.05% among individuals 19 years of age and younger, between 0.19% and 2.7% for individuals between 20-59 years old, and is at least 9.5% for individuals older than 60. Because estimating the number of deaths is not a primary aim of this model, however, uncertainty in the mortality risk is unlikely to have a significant impact on the model results. The main effect of the uncertainty would be to slightly change the size of the recovered population, which would impact the rate of exposure for susceptible individuals, but likely not enough to affect the main conclusions of the analysis.

#### 2.2 Calibrated Parameters

**Table 2:**
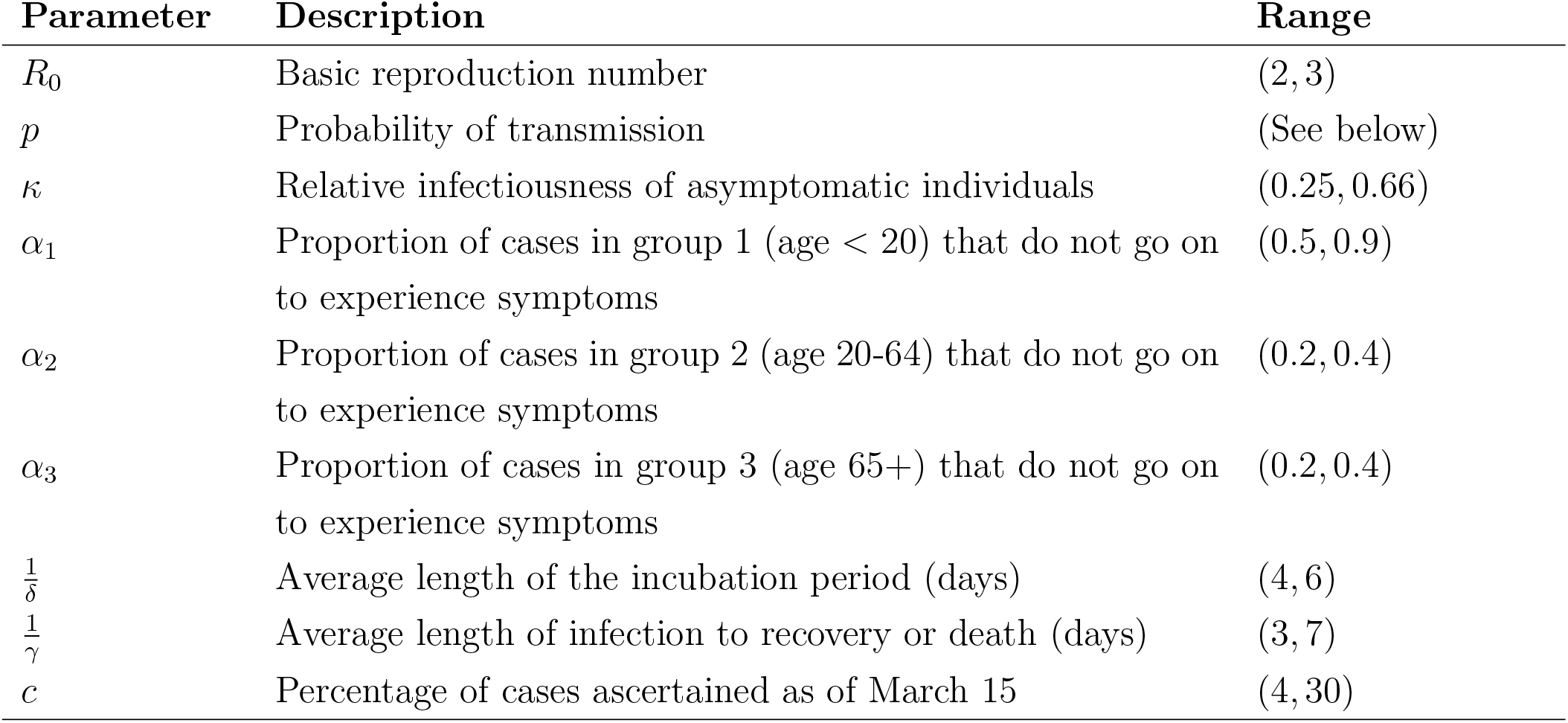
Calibrated Parameters

##### 2.2.1 Basic Reproduction Number

Both Li et al. (2020a) and Riou and Althaus (2020) (90% high density interval: 1.4–3.8) estimate *R*_0_ at 2.2, with Li et al. (2020a) estimating a 95% confidence interval of (1.4, 3.9) and Riou and Althaus (2020) estimating a 90% high density interval of (1.4,3.8). Other models have assumed an *R*_0_ around this 2.2 value, but with varying levels of uncertainty: using Li et al. (2020a) and Riou and Althaus (2020), Peak et al. (2020) assume a 95% confidence interval of (1.46, 3.31), Ferguson et al. (2020) examines values between 2.0 and 2.6, and Prem et al. (2020) adapts a model from Kucharski et al. (2020) to estimate an interquartile range of (1.6, 3.0). In line with this general range of uncertainty, we calibrate *R*_0_ over a range from 2 to 3.

##### 2.2.2 Probability of Transmission

There is no direct data available on the probability of transmission per contact, so the value of this parameter was set as a function of *R*_0_. A value for *p* was calibrated to values of *R*_0_ from 2 to 3: the model was run with the baseline values for the other parameters listed in Tables 1 and 3, and *p* was adjusted so that the model output fit each target value of *R*_0_. Then, in the main calibration procedure described below, *p* was determined by the *R*_0_ drawn for each parameter set, based on this initial mapping between *p* and *R*_0_ values.

##### 2.2.3 Relative infectiousness of asymptomatic individuals

We assume that the rate of transmission from asymptomatic individuals is systematically lower than the transmission probability from symptomatic individuals, but there are few empirical estimates for this reduction in transmission. We calibrated the relative infectiousness of asymptomatic individuals across a range of 25% to 66%. This range was derived from the values used by other researchers: Zhao et al. (2020) used 50% (which is similar to the estimate for influenza); Prem et al. (2020) used 25% (which they derived from Liu et al. (2020)); and Ferguson et al. (2020) used 66%.

##### 2.2.4 Proportion of asymptomatic infections

Mizumoto et al. (2020) estimate that, among individuals on the Diamond Princess Cruise ship, 20.6% to 39.9% of infected individuals are asymptomatic, depending on the mean incubation period used by their model (they evaluated mean incubation periods between 5.5 and 9.5 days). Based on these results, we calibrate the proportion of asymptomatic infections for individuals 20 years and older across the range of 20% to 40%. This range is also supported by Nishiura et al. (2020), which found that 30.8% (95% CI: 7.7, 53.8) of infected individuals among Japanese citizens evacuated from Wuhan, China were asymptomatic.

There is less available data on the proportion of asymptomatic infections among individuals younger than 20 years, Russell et al. (2020) find that, among 6 cases detected in individuals under 20 years old as of Februrary 20, 2020 on the Diamond Princess Cruise ship, 4 of the cases were asymptomatic. Therefore, we calibrate across a wide range around this naive estimate of 66%, from 50% to 90%.

##### 2.2.5 Incubation and Latent period

Li et al. (2020a) reported a mean incubation period of 5.2 days in the first 425 confirmed patients from Wuhan, China; Linton et al. (2020) estimate a mean period of 5.6 days for patients in China (both inside and outside Wuhan); Lauer et al. (2020) estimates the median incubation period at 5.1 days for patients outside Hubei province, China; while Xu et al. (2020) reports a slightly lower median period of 4 days for patients in Zhejiang province, China, and Backer et al. (2020) reports a slightly higher mean period of 6.4 days for travelers from Wuhan, China. Based on these results, we calibrate the length of the incubation period across a range of 4 to 6 days.

Additionally, it is important to note that we assume the incubation and latent periods coincide and that there is no pre-symptomatic infectious period. This is likely not an accurate assumption, given reports of pre-symptomatic transmission, but unfortunately no data is available on the length of the latent period itself. This assumption will be revisited and updated in the model as more data becomes available.

##### 2.2.6 Duration of Infectious Period

There is limited evidence available on the duration of the infectious period. Prem et al. (2020) models a length of both 3 and 7 days, so we calibrate our model across this range.

##### 2.2.7 Proportion of cases detected

Because of the lack of widespread testing, there is major uncertainty around the proportion of cases detected. McAndrew and Reich (2020) conducted an expert survey that estimated only 13% of all cases have been detected, with an 80% uncertainty interval of 4-30% – we calibrate the parameter value in our model across this range. This range is further supported by recent research from Li et al. (2020b), which estimates that 86% of all cases are undetected.

### 3 Calibration Procedure

The parameters listed in the section above were calibrated using a Bayesian approach to estimate the posterior distribution for these parameters, operationalized using sampling importance resampling (SIR) (Raftery and Bao, 2010). The target data comprised of confirmed COVID-19 cases through March 15, 2020, across six counties in the San Francisco Bay Area.

To implement SIR, we drew 10,000 vectors of parameters, with each parameter drawn from an independent uniform distribution spanning the range in Table 2. For parameters not listed in Table 2, we used base case values specified in Table 1. We then computed the likelihood of each parameter vector given the target data, assuming that cumulative cases on March 15 were distributed normally and using the drawn value of *c* to determine the number of known cases estimated by the model. Finally, we resampled parameter vectors from the original set of 10,000, with replacement, using sampling probabilities proportional to the computed likelihood of each parameter vector in order to obtain an analytical estimate of the posterior distribution given the prior and our model.

### 4 Estimation of Hospitalizations

We estimated hospitalizations through an application of Bayes’ rule. Let *T* be the event that a COVID-19 patient has a positive test and *H* be the event that a COVID-19 patient is hospitalized. Then, the unconditional probability that a COVID-19 patient is hospitalized is:

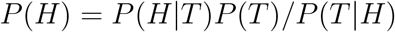

We assume that *P* (*H*|*T*) = 0.33 based on an approximation from *Coronavirus (COVID-19) Data Dashboard - Public Health Department - County of Santa Clara* (n.d.), *P* (*T*) = *c* (see above), and *P* (*T* |*H*) *∼* 1, based on California’s testing policies during the calibration period. To calculate the number of hospitalized cases at each time point, we assumed that patients stay an average of 10 days in the hospital, following Ferguson et al. (2020). We obtained capacity estimates for 6 counties based on the American Hospital Association Survey and assumed that 50% of beds (*Coronavirus (COVID-19) Data Dashboard - Public Health Department - County of Santa Clara* (n.d.), approximation) can be made available to COVID-19 patients: a total of 2271 beds for our capacity constraint (American Hospital Association, 2018).

